# Effects of a single 10mg dose of empagliflozin on postprandial insulin kinetics in patients with postbariatric hypoglycaemia

**DOI:** 10.1101/2021.11.01.21265679

**Authors:** Michele Schiavon, David Herzig, Hepprich Matthias, Marc Y. Donath, Chiarra Dall Man, Lia Bally

## Abstract

**Introduction:** Postbariatric hypoglycaemia (PBH) is an increasingly recognized late metabolic complication of Roux-en-Y gastric bypass (GB) surgery. PBH typically manifests with a fact occurring post-meal hyperglycaemic peak, followed by a disproportionately exaggerated insulin response leading to low glucose levels. On this basis, we evaluated the effect of a single dose of empagliflozin 10mg vs. placebo on parameters of insulin kinetics.

**Materials and methods:** Insulin secretion, hepatic insulin extraction and total insulin clearance were evaluated after a single of empagliflozin 10mg vs. placebo followed by a standardized liquid mixed meal were evaluated in 11 subjects with confirmed PBH after GB over 3h. Parameters of interest were calculated using established mathematical models. Indices were compared between the groups using the Wilcoxon signed-rank test.

**Results:** Total beta-cell responsiveness tends to be lower with empagliflozin vs. placebo (24.83±11.00 vs. 27.15±9.68 [10^−9^ min^-1^], p=0.150). Total first-pass hepatic insulin extraction increased after empagliflozin compared to placebo (49.6±14.2 vs. 39.7±12.1 %, p=0.006), while no significant effect of empaglizflozin on basal first-pass hepatic insulin extraction was observed (79.7±7.1 vs. 81.1±6.6 %, p=0.521). Total insulin clearance resulted to be significantly lower after empagliflozin compared to placebo (3.91±1.58 vs. 3.00±1.27 l/min, p=0.002).

**Conclusion:** The present analysis suggests that the hypoglycaemia-attenuating effect of SGLT2-inhibition in patients with PBH is mainly mediated by an increment in insulin clearance, with also a tendency to a reduction in insulin secretion.

## INTRODUCTION

Roux-en-Y gastric surgery (GB) is one of the most effective anti-obesity treatments, but is however not free from complications(*1*). One major complication is postbariatric hypoglycaemia (PBH) – occurring in up to 30% of patients. The hypoglycaemia typically occurs in the late postprandial period, after a fast occurring hyperglycaemic peak, followed by a disproportionally exaggerated insulin response.

Despite the potentially deleterious consequences of PBH, there are currently no approved pharmacotherapies for this condition. In the Delphi consensus published in 2020(*2*), acarbose, a glucosidase inhibitor that lowers postprandial glucose excursion is recommended to treat PBH with a level B evidence. However, the inconvenience of administration (before every meal) and the frequent side effects (flatulence due to carbohydrate malabsorption) results in poor uptake of this medication.

A pharmacological alternative to acarbose that similarly lowers postprandial glycaemia, albeit by a different mechanism, are inhibitors of the sodium glucose co-transporter 2 (SGLT2-inhibitors) which mediate glucose re-absorption in the kidney(*3*). Inhibition of SGLT2 decreases renal glucose reabsorption, promotes urinary glucose excretion and reduces plasma glucose concentrations through an insulin-idependent mechanisms.

Recently, a single 10mg dose of SGLT2-inhibitor empagliflozin was shown to significantly reduce the postprandial hypoglycaemia burden during a mixed meal tolerance test in patients with PBH(*4*). Compared to placebo, lower hypoglycaemia burden was accompanied by significantly decreased insulin exposure.

In this subanalysis, we investigate the underlying mechanisms by which empagliflozin reduces insulin exposure in patients with PBH, namely its effects on insulin secretion (e.g. basal and postprandial beta-cell glucose responsiveness) and insulin clearance (e.g., first pass hepatic insulin extraction and total insulin clearance).

## METHODS

### Clinical experiments

This was a retrospective analysis on data from a randomized control trial comparing the efficacy of empagliflozin vs. placebo to reduced postprandial hypoglycaemia in PBH (*5*). Twelve post-GB adults with PBH (documented plasma glucose ≤2.5mmol/L with symptoms) received a single dose of empagliflozin (10mg) or placebo 2h before a standardized liquid mixed-meal containing 60g of carbohydrates and underwent frequent blood sampling for up to 3h. Protocols were approved by the local Ethics Committee. Participants provided written informed consent and the protocol was approved the local Ethics Committee Basel.

### Laboratory analyses

Blood glucose concentration was quantified by the Contour XT point-of-care meter (Ascensia, Diabetes Care, Switzerland). Commercial immunoassays were used to determine insulin (Elecsys Insulin, Cobas, Roche Diagnostics, Mannheim, Germany) and C-peptide (Mercodia AB, Uppsala, Sweden) concentrations.

### Mathematical modelling of parameters regulating insulin exposure

Beta-cell glucose responsiveness and first pass hepatic insulin extraction were estimated by means of the Oral C-peptide Minimal Model (OCMM) coupled with a model of insulin extraction and kinetics (IMM). The OCMM and the IMM were used to describe C-peptide and insulin data and estimate C-peptide secretion and first-pass hepatic insulin extraction. C-peptide and post-hepatic insulin kinetics (required for the OCMM and IMM identification) were estimated from the data using an extended version of a recently proposed Bayesian approach (*6*), but these were assumed to be identical on both visits helping to avoid possible compensations against beta-cell responsivity and hepatic insulin extraction. Additionally, we also estimated total insulin clearance (composite of hepatic and post-hepatic elimination) using a formula reported in Gastaldelli et al. (*7*).

### Statistical analysis

Indices were compared between the conditions using paired T-tests. Normality assumption was visually assessed using quantile-quantile plots. Results are reported as mean±standard deviation (SD) unless otherwise specified. P-values <0.05 were considered as statistically significant. Statistical analysis were performed with R (version 4.0.2).

## RESULTS

Data from 11 participants with PBH (9F/3M; age=44±10y; BMI=27.4±3.9kg/m^2^) were included in the analysis. A single dose of empagliflozin 10mg compared to placebo lowered postprandial plasma insulin exposure by 30±21%. Reduced insulin exposure was attributable to both reduced insulin secretion and, increased insulin clearance, as outlined below.

### Effect on insulin secretion

Total beta-cell responsiveness [ϕ_tot_], which represents the overall ability, i.e. combining basal [ϕ_b_] and over-basal [ϕ_ob_] contributions, of glucose to stimulate beta-cell independent of insulin sensitivity tends to be lower with empagliflozin vs. placebo (24.83±11.00 vs. 27.15±9.68 [10^−9^ min^-1^], p=0.150). In particular, basal beta-cell responsiveness [ϕ_b_] resulted to be significantly reduced by empagliflozin (6.85±2.67 vs. 8.30±3.50 10^−9^min^-1^, p=0.0019), while over-basal beta-cell responsiveness tends to be higher [ϕ_ob_] (125.70±80.28 vs. 103.83±54.22 10^−9^min^-1^, p=0.055).

### Effect on insulin clearance

Total first-pass hepatic insulin extraction increased after empagliflozin compared to placebo (49.6±14.2 vs. 39.7±12.1 %, p=0.006), while no significant effect of empaglizflozin on basal first-pass hepatic insulin extraction was observed (79.7±7.1 vs. 81.1±6.6 %, p=0.521). Total insulin clearance, as calculated from (*7*), resulted to be significantly lower after empagliflozin compared to placebo (3.91±1.58 vs. 3.00±1.27 l/min, p=0.002).

## DISCUSSION

The present work provides evidence that a single dose of empagliflozin 10mg significantly reduces insulin exposure in individuals suffering from PBH after GB surgery, mainly by an increment in insulin clearance while only a tendency in reducing insulin secretion, when compared to placebo. Of note, beta-cell glucose sensitivity as a functional parameter of insulin secretion was only reduced in the fasted but not during the postprandial status. The enhancing effect of empagliflozin on insulin clearance, when assuming post-hepatic insulin clearance to be the same of placebo, was demonstrated by a significant increase in total first pass hepatic insulin extraction.

Inadequate insulin exposure appears to be a key pathophysiological feature of PBH which is a increasingly recognized late metabolic complication of GB with potentially debilitating consequence. Only recently, the efficacy of a single dose of 10mg empagliflozin in reducing the burden of PBH after a mixed-meal was shown for the first time (*4*). Two weeks of an alternative SGLT2-inhibitor, canagliflozin in a dose of 300g, was also recently shown to prevent PBH following an oral glucose load (*8*). However, canagliflozin, unlike empagliflozin, also inhibits the intestinal SGLT1 (*9*) with impact on both postprandial glucose excursion and intestinal peptide responses. Thus, the underlying mechanisms by which hypoglycaemia protecting effects are exerted differ between the two agents. Nonetheless these first insights allude to the promising effects of SGLT2-inhibitor as well-tolerable and efficacious treatment of PBH. We are therefore conducting a randomised controlled trial to evaluate the potential of empagliflozin in lowering the burden of PBH in an unrestrained outpatient setting (NCT05057819).

Consistent with its importance in determining plasma insulin levels, insulin clearance seems to be substantially influenced by empagliflozin. At this stage, the interpretation of this finding remains speculative. As high portal insulin concentrations have been suggested to be associated with decreased insulin clearance due to receptor saturation(*10, 11*), the represent findings may be seen in the light empagliflozin-induced lowering of insulin secretion. However, not all studies support reduced fractional hepatic insulin extraction with increasing glucose loads(*12*). Of note several studies reported a link between total insulin clearance and whole body insulin sensitivity(*13*). Although it is conceivable that empagliflozin influences insulin action, the dataset used for the analysis did not allow us to quantify insulin sensitivity and its relationship with other outcomes as glucosuria was not quantified in the experiment.

The strength of this work is the mechanistic exploration of empagliflozin in the context of PBH, an emerging condition with comparably high prevalence and clinical relevance. However, we acknowledge several limitations. First, the data for the analyses were collected after a single dose regimen which is not representative of a steady-state condition. Second, the effect on urinary glucose excretion which mainly occurs two hours post-ingestion was not obtained and thus precluded the calculation of non-insulin and insulin-dependent glucose disposal. Third, although the used models enabled measures of hepatic insulin extraction to be estimated, the fractional extraction calculation was based only on newly secreted insulin, which represents only a portion of the insulin delivered to the liver, while the rest coming from the recirculation of plasma insulin through the portal vein and hepatic artery, which in many conditions is greater than the endogenous secretion rate. Although we additionally estimated total insulin clearance by a different approach, relative contributions of hepatic and post-hepatic clearance cannot be clearly distinguinshed in the current dataset. In fact, in our modelling analysis no improvements in model prediction was observed when estimating both hepatic and post-hepatic insulin clearance (not shown), hence we assumed the latter to be the same between visits. However, the conductance of extra experiments are needed to better understand their relative contributions. And fourth, empagliflozin may impact on other gluco-regulatory factors (e.g. glucagon, GLP-1) with potentially hypoglycaemia-protecting effects(*14*). However, glucagon which acts as the most imminent insulin counter-regulatory hormone and the insulinotropic GLP-1 remained unchanged after a single 10mg dose of empagliflozin(*5*).

In conclusion, the present analysis suggests that the hypoglycaemia-attenuating effect of SGLT2-inhibition in patients with PBH is mainly mediated by an increment in insulin clearance, with also a tendency to a reduction in insulin secretion. Ongoing longer studies using empagliflozin in PBH patients will unravel whether its regulatory effects on insulin exposure and potentially other relevant factors will prove its worth in clinical practice.

## Data Availability

All data produced in the present work are available upon reasonable request to the authors

## Acknowledgements

The authors thank all study participants for their time and efforts.

## Author contributions

LB, CDM, DH and MS designed the retrospective analysis. MH and MYD provided the data. MS analysed the data. LB, CDM, DH and MS supported the analysis, interpreted the data and wrote the manuscript. All authors critically reviewed the manuscript. LB is the guarantor of this work and takes responsibility for the integrity of the data and the accuracy of the data analysis.

